# Plasma proteomics link menopause timing to brain aging and dementia risk

**DOI:** 10.64898/2026.04.23.26351500

**Authors:** Madeline Wood Alexander, Brendan Wood, Hamilton See-Hwee Oh, Veronica Augustina Bot, Julia Borger, Francesca Galbiati, Keenan A. Walker, Susan M. Resnick, Heather M. Ochs-Balcom, Tony Wyss-Coray, Charles Kooperberg, Alexander P. Reiner, Emily G. Jacobs, Jennifer S. Rabin, Kaitlin B. Casaletto, Rowan Saloner

## Abstract

Earlier menopause is a risk factor for several age-related diseases, including dementia. The biological pathways linking menopause timing to later-life brain aging are not understood. Leveraging large-scale plasma proteomics in postmenopausal women from the UK Biobank (*N*=15,012), earlier menopause was associated with upregulation of pro-inflammatory and extracellular matrix degradation pathways, plus accelerated aging across proteomic clocks of organ and cellular aging, including brain and oligodendrocyte aging. Elevated GDF15, a canonical aging marker, was the top protein correlate of earlier menopause. We observed robust replication of menopause timing proteomic shifts in the Women’s Health Initiative Long Life Study (*N*=1,210). In UKB, proteins associated with earlier menopause, including GDF15, exhibited concordant associations with incident dementia risk and brain atrophy, cerebral small vessel disease burden, and white matter microstructural integrity. Collectively, our findings identify proteomic signatures linking ovarian aging to brain aging, providing a framework to inform interventions to reduce dementia risk.

## INTRODUCTION

Menopause is a critical transition in female biological aging with widespread implications for disease risk.^1–8^ Underpinned by ovarian aging, which occurs earlier and more rapidly than aging in other organs, menopause is marked by a dramatic endocrine shift that affects multiple physiological systems including the brain.^1–3,9–11^ Emerging evidence suggests that hypothalamic aging may precede ovarian senescence and trigger menopause timing,^12,13^ directly implicating the brain in the menopause cascade. Menopause occurs in the coincident midlife window of when the earliest pathological manifestations of neurodegenerative diseases incite^14,15^ and may contribute to well-established sex differences in risk and resilience to dementia.^4,16–20^

Earlier menopause is associated with accelerated cognitive aging and increased risk for dementia.^5,6,21–23^ The mechanisms linking menopause timing to greater dementia risk are unclear, but some studies indicate that women with earlier menopause show enhanced susceptibility to multiple dementia risk factors, including elevations in both AD and non-AD pathology, cerebrovascular burden, and synaptic dysfunction.^5,6,8,22^ In addition to dementia, earlier menopause is associated with increased risk for a range of age-related diseases,^24–27^ thereby connecting earlier ovarian aging to accelerated biological aging across organ systems.^28^

Menopause affects half of the global population, yet we lack a fundamental understanding of how this universal female transition influences the brain. Recent large-scale proteomics studies have identified molecular signatures of brain aging and neurodegenerative diseases enriched for immune, metabolic, synaptic, and vascular biology.^29–33^ These systems are also altered during menopause,^1–3,9–11^ and may represent pathways connecting age at menopause to dementia risk. However, the blood-based proteomic profiles of menopause timing have not yet been characterized in human cohorts or linked to the aging female brain.

Using plasma proteomics data from two large independent cohorts, we characterized and validated biological signatures of menopause timing, including proteomic alterations to inflammatory, metabolic, extracellular matrix remodeling, and apoptotic pathways. We demonstrate that earlier menopause is associated with accelerated biological aging across proteomic clocks of most organ systems and cell types, including the brain. We observed that proteomic shifts tied to earlier menopause also predicted future risk for dementia and tracked with concurrent magnetic resonance imaging (MRI) measures of brain volume, small vessel disease burden, and white matter integrity. Our findings provide a comprehensive and validated proteomic signature of menopause timing that captures the molecular overlap between earlier menopause and accelerated brain aging, revealing novel pathways that may underlie dementia risk.

## RESULTS

### The plasma proteomic signatures of menopause timing

The UK Biobank (UKB) discovery cohort included *N*=15,012 postmenopausal women with Olink proteomics who were an average of 10.5 (*SD*=6.6) years post-menopause (mean[*SD*] age=60.7[5.07], mean[*SD*] age at menopause=50.1[4.65]; Tab.S1). In all analyses, age at menopause was modelled continuously. Linear models adjusted for chronological age showed that age at menopause was positively associated with 105 proteins and negatively associated with 563 proteins (Fig. 1a-b; Tab.S2). Top hits upregulated with earlier age at menopause included immune markers GDF15 and LAMP3. Top hits upregulated with later age at menopause included bone marker OMD, extracellular growth factor regulators WFIKKN1 and WFIKKN2, and lipid regulator PM20D1. Gene ontology (GO) and pathway enrichment analyses revealed that earlier menopause associated with increases in proteins related to immune activation, apoptosis, and extracellular matrix remodeling. Conversely, later age at menopause associated with increases in proteins related to bone maintenance, lipid transport, and growth factor signaling (Fig. 1c).

**Figure 1.**
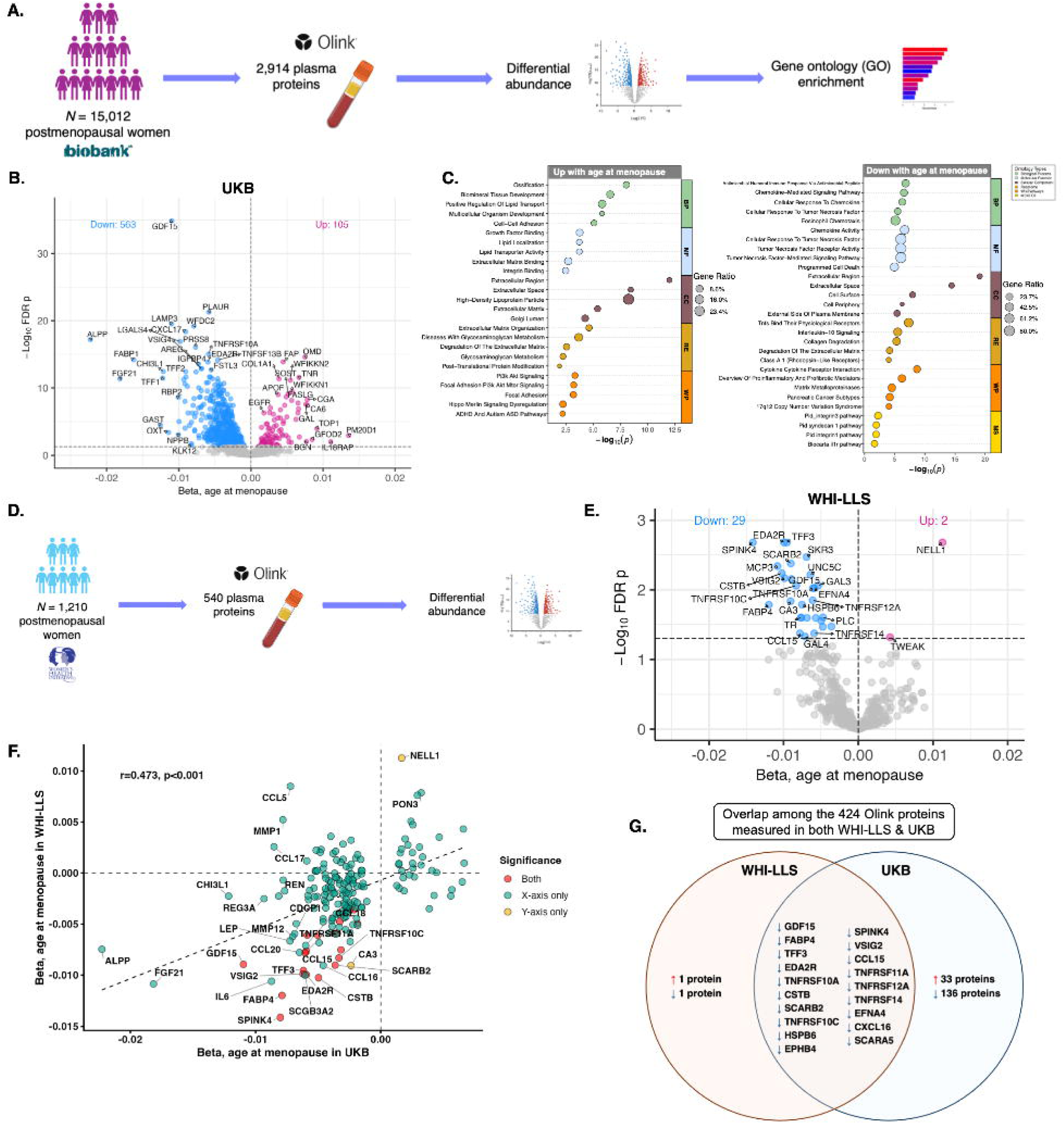
Plasma proteomic signatures of menopause timing. **A. Discovery cohort overview.** Main analyses included *N*=15,012 postmenopausal women ≥50 years old with 2,916 Olink plasma proteins in the UK Biobank (UKB) discovery cohort. **B. Differential abundance by age at menopause in UKB.** Age-adjusted linear models revealed that age at menopause was positively associated with 105 proteins and negatively associated with 563 proteins (FDR *p*<.05). **C. Gene ontology (GO) and pathway enrichment for menopause timing proteins.** Age at menopause was associated with upregulation in proteins related to bone health, growth factor, metabolism, and survival activity, and downregulation in proteins related to inflammation, apoptosis, extracellular matrix remodeling and repair. **D. Validation cohort overview.** We validated menopause timing protein associations in *N*=1,210 postmenopausal women ≥65 years old with 540 Olink plasma proteins in the Women’s Health Initiative (WHI) Long Life Study (LLS) Visit 1.**E. Menopause timing proteomic shifts replicate cross-cohort.** In the WHI-LLS sample, adjusted for age and original hormone therapy randomization status, age at menopause was positively associated with levels of 2 proteins, and negatively associated with levels of 29 proteins (FDR *p*<.05). **F. Comparison of menopause timing protein effect sizes between the discovery cohort (UKB) and validation cohort (WHI-LLS).** Among the 424 Olink proteins available in both UKB and WHI-LLS, there was strong concordance of the effect sizes for the associations between age at menopause and protein levels. **G. Overlap between UKB and WHI-LLS menopause proteins.** 19 proteins significantly (FDR) negatively associated with age at menopause in both cohorts.

We next validated the UKB findings in the Women’s Health Initiative (WHI) Long Life Study (LLS) by examining associations between age at menopause and plasma Olink proteins in *N*=1,210 postmenopausal women (mean[*SD*] age=80.3[6.39], mean[*SD*] age at menopause=48.1[6.61]; Tab.S3) in linear models adjusted for age and original WHI randomization status (i.e., hormone therapy vs. placebo/observational; Fig. 1d). We observed strong concordance (r=0.47) for menopause timing effect sizes across the 424 plasma proteins available in both the UKB and WHI-LLS datasets (Figs. 1e-f; Tab.S4). The WHI-LLS proteomics had greater coverage of proteins negatively associated with age of menopause, including 19 proteins that reached false discovery rate (FDR) significance in both UKB and WHI-LLS (Fig. 1g). Top replicated hits included several inflammatory and tissue injury-related proteins, including GDF15, as well as epithelial repair protein SPINK4, adipokine FABP4, and multiple members of the TNF receptor superfamily (e.g., TNFRSF10C, TNFRSF11A). Given that the WHI-LLS is a multiethnic cohort (Tab. S3) while UKB is predominantly White/British (Tab.S1), we also considered analyses adjusted for race and ethnicity (Tab.S5), which yielded very similar findings to the primary WHI-LLS analyses (r=0.95; Fig S1), as well as consistent results with both primary UKB analyses (r=0.48) and race/ethnicity-adjusted models in UKB (r=0.47; Tab.S6). These results provide strong replication of key menopause timing-related proteomic shifts across independent cohorts of postmenopausal women of different age and demographic groups.

### Earlier menopause associates with proteomic signatures of accelerated biological aging

We evaluated the associations of age at menopause with measures of proteomic aging in the UKB, using previously established proteomic clocks for 13 organ systems and 38 cell types (Fig. 2a).^34,35^ For organ aging, age-adjusted linear models showed that earlier age at menopause associated with larger (i.e., less favourable) age gaps on measures of brain, heart, immune [blood/spleen], liver, pancreas, lung, kidney, adipose, and intestine aging, plus larger age gaps on measures of overall organ-shared proteomic aging (i.e., conventional, organismal; Fig. 2b; Tab.S7). Menopause timing did not significantly associate with artery or muscle aging. A chord plot shows top menopause proteins from the most statistically significant (FDR-*p*) menopause timing-associated measures of organ aging (Fig. 2c). A visualization of LOESS-estimated slopes of brain aging proteins associated with menopause timing showed upregulation of neuronal extracellular matrix structure and signaling proteins with later menopause (e.g., TNR, BCAN, MEPE, CNTN1) and upregulation of core markers of neural and glial injury with earlier menopause (e.g., NEFL, MOG, SNAP25, GFAP; Fig. 2d).

**Figure 2.**
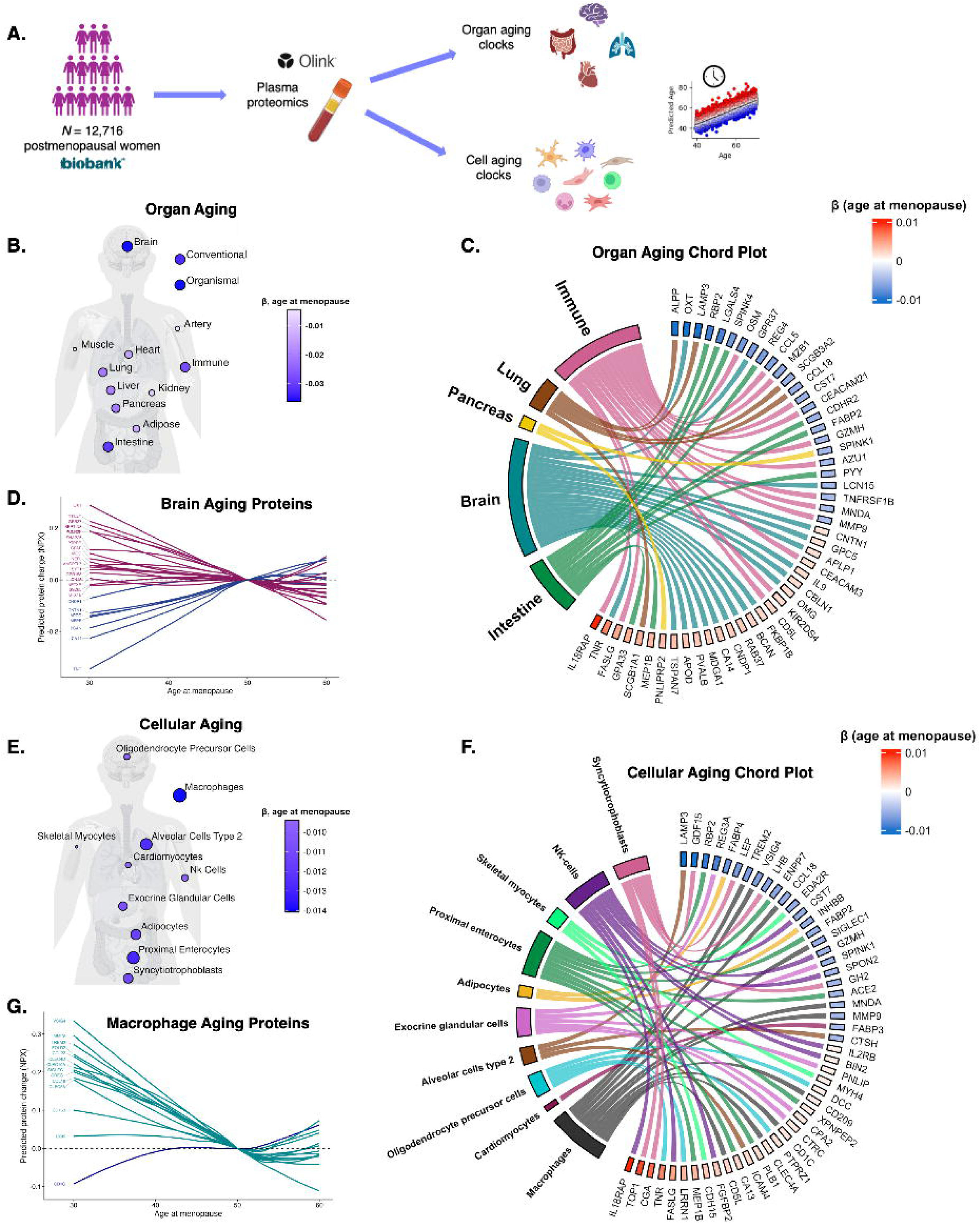
Earlier menopause associates with multi-system accelerated biological aging. **A. Overview.** In *N*=12,716 postmenopausal women ≥50 years old in the UK Biobank (UKB), we estimated 13 plasma proteomic clock measures of organ-specific biological aging. **B. Age at menopause associates with proteomic clocks of organ aging.** The body plot shows that older age at menopause was associated with smaller age gaps on models predicting biological age across most organ systems (FDR *p*<.05). **C. Top menopause timing proteins from from intestine, brain, pancreas, lung, and immune systems.** Among proteins that are enriched in each of the top five most significant menopause timing-associated organ age measures, the chord plot shows the 25 proteins that positively associate with age at menopause and the top 25 proteins that negatively associate with age at menopause. **D. Predicted changes in brain aging proteins by age at menopause.** 24 brain aging proteins also significantly associated with age at menopause (FDR *p*<.05). For each of these proteins, the plot shows the predicted change in level (NPX) by age at menopause. Estimates were extracted from age-adjusted LOESS regression models, and trajectories were centered at age at menopause = 50. Blue lines are trajectories for proteins that were positively associated with age at menopause in linear fits, and purple lines are trajectories for proteins that were negatively associated with age at menopause in linear fits. **E. Age at menopause associates with proteomic clocks of cell aging.** Older age at menopause associated with smaller age gaps for 35 of the 39 cellular aging clocks examined. The body plot shows the top ten strongest menopause-timing associated cell clocks (smallest FDR *p*s. **F. Top menopause timing proteins originating from intestine, brain, pancreas, lung, and immune systems.** Among proteins that are enriched in the ten most significant menopause timing-associated cell age measures, the chord plot shows the 25 proteins that positively associate with age at menopause and the top 25 proteins that negatively associate with age at menopause. **G. Predicted changes in macrophage aging proteins by age at menopause.** 14 macrophage aging proteins also significantly associated with age at menopause (FDR *p*<.05). For each of these proteins, the plot shows the predicted change in level (NPX) by age at menopause. Estimates were extracted from age-adjusted LOESS regression models, and trajectories were centered at age at menopause = 50. Turquoise lines are trajectories for proteins that were positively associated with age at menopause in linear fits, and navy lines are trajectories for proteins that were negatively associated with age at menopause in linear fits.

Regarding cellular aging, age-adjusted linear models revealed that earlier menopause was associated with larger age gaps on 33 of the 38 cell clocks examined (Tab.S8, Fig S2). The strongest associations with menopause timing (i.e., smallest FDR-*p*) were observed for aging measures for macrophages, proximal enterocytes, syncytiotrophoblasts, and alveolar cells (Fig. 2e). Among brain cells, glial clocks exhibited stronger associations with menopause timing than neuronal clocks (Tab. S8). The largest glial cell clock association with menopause timing was for oligodendrocyte precursor cell aging, which was previously shown to exhibit the strongest association with dementia severity in neurodegenerative diseases.^35^ A chord plot showed that age at menopause was positively associated with oligodendrocyte precursor cell proteins implicated in axon guidance and neuroplasticity (i.e., TNR, LRRN1, PTPRZ1, DCC; Fig. 2f). LOWESS-estimated slopes of macrophage aging proteins (the strongest cell aging correlate of menopause timing) revealed increases in a range of pro-inflammatory proteins with earlier menopause, including VSIG4, MMP-9, and TREM2 (the latter of which is implicated in AD;^36^ Fig. 2g). VSIG4 is upregulated in ovarian cancer,^37^ which has previously been linked to earlier menopause.^38,39^

Earlier menopause also associated with accelerated syncytiotrophoblast aging. Postmenopausal women do not harbor these placental-derived cells, but many of the syncytiotrophoblast aging markers are implicated in pituitary- and hypothalamic-mediated reproductive and endocrine pathways (e.g., CGA, LHB, LEP, GDF15; Fig. 2f), such that aging on this measure may proxy aging of hormone responsive biology more broadly. Thus, we next examined whether menopause timing proteins were enriched in other endocrine-regulated female tissues, which could include markers that comprised the syncytiotrophoblast aging signature, as well as other female cell type-specific proteins significantly associated with menopause timing (e.g., oocyte aging proteins; Tab.S8). Specifically, we investigated whether menopause timing proteins were disproportionately represented by targets with high expression in female reproductive tissues (i.e., endometrium, ovary, fallopian tube, cervix, vagina, breast, placenta) using Fisher’s exact tests. Female reproductive tissue proteins were disproportionately represented among targets negatively associated with age at menopause (OR=1.81, *p*=.04), including targets implicated in endocrine and metabolic processes such as the cervix- and placenta-enriched metalloenzyme ALPP, breast-enriched LEP, and the cervix- and vagina-enriched serine protease KLK12 (Tab.S9). Notably, none of the proteins positively associated with age at menopause were female reproductive tissue enriched.

### Proteomic signatures of earlier menopause are distinct from those of surgical menopause and menopause stage

To examine whether the age of menopause proteomic profile was influenced by overrepresentation of surgical menopause (i.e., bilateral oophorectomy before menstrual cessation) among women with earlier menopause, we next evaluated associations of menopause type (surgical [*N*=397] vs. spontaneous/natural [*N*=14,407]) with protein levels in multivariable models adjusted for age at menopause, chronological age, and history of hormone therapy. After accounting for these factors, surgical menopause was associated with upregulation of 55 proteins and downregulation of 33 proteins (Fig.S3; Tab.S10). ENPP2 (i.e., autotaxin), an immune-activated lipid metabolism enzyme expressed in uterine tissues,^40^ was the top upregulated protein associated with surgical menopause. Top downregulated proteins included ovary-expressed protein WFIKKN2 and endocrine and neuroendocrine protein CGA, reinforcing the role of ovarian and reproductive endocrine axis signaling even after menopause.^1^ WFIKKN2 and CGA were among a small number of proteins that exhibited overlapping significant associations with both menopause type and timing (*N*=6 overlapping proteins). There was minimal concordance between menopause type and menopause timing effect sizes on the proteome (r=-0.016, Fig. 3a).

**Figure 3.**
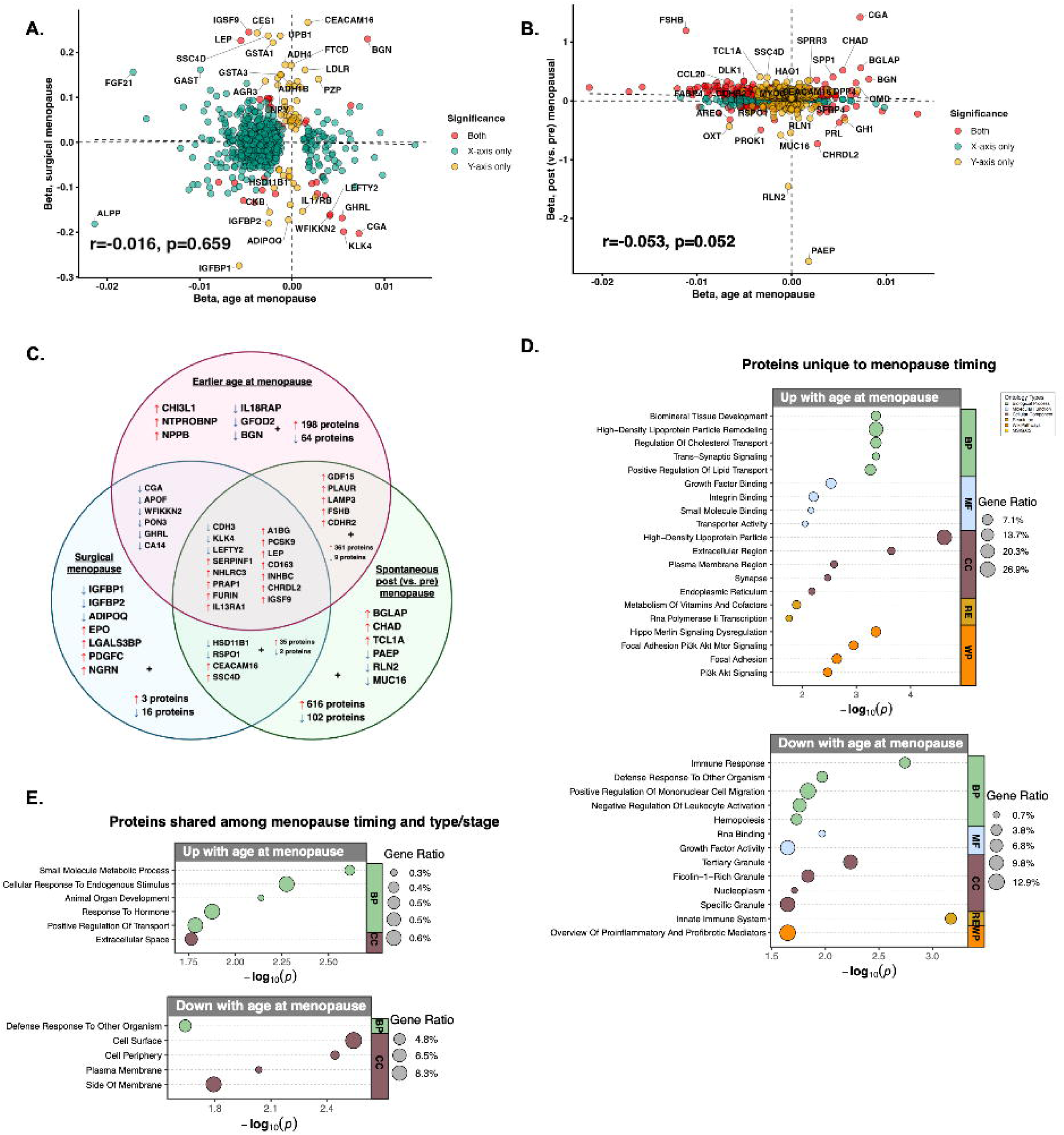
Plasma proteomic profiles of earlier menopause show differences from those of menopause type and menopause stage. **A. Comparison of protein effect sizes for age at menopause versus menopause type.** There was minimal concordance between protein associations with age at menopause (adjusting for menopause type, age, and hormone therapy) and protein associations with surgical menopause (adjusting for age at menopause, age, and hormone therapy). **B. Comparison of protein effect sizes for age at menopause versus menopause stage.** There was minimal concordance between protein associations with age at menopause (adjusting for menopause type, age, and hormone therapy) and protein associations with menopause stage, using summary statistics from a previous analysis of age-matched post- and premenopausal women in the UKB, suggesting that earlier menopause and the menopause transition itself have unique proteomic signatures. **C. Venn diagram comparing the proteomic correlates of earlier (vs. later) age at menopause, surgical (vs. spontaneous) menopause, and post (vs. pre) menopausal status.** 15 proteins showed directionally concordant and statistically significant associations with all three menopause-related factors. **D-E. Gene ontology (GO) and pathway enrichment.** GO enrichment terms for proteins unique to menopause timing (D) exhibit distinct biology from those shared among menopause timing proteins and either menopause stage and/or menopause type.

To disentangle effects of menopause stage from menopause timing, we also compared menopause timing proteins against summary statistics describing differential protein abundance in age-matched pre- and postmenopausal women in the UKB (*N*=2,814; aged 45-60, mean[*SD*] age premenopausal=50.9[2.51], mean[*SD*] age postmenopausal=50.9[2.62]).^20^ There were 375 proteins that showed statistically significant and directionally concordant associations between menopause timing and stage. These included 366 proteins that were both negatively associated with age at menopause and upregulated in post (vs. pre) menopausal women, including the sex hormone and menopause biomarker FSHB, and 9 proteins that were both positively associated with age at menopause and downregulated in post (vs. pre) menopausal women, including the pituitary protein prolactin (PRL). However, there was minimal correlation between effect sizes for protein associations with menopause timing and menopause stage (r=-0.053, Fig. 3b).

Between earlier (vs. later) age at menopause, surgical (vs. spontaneous) menopause, and post (vs. pre) menopausal status, there were 15 proteins that showed directionally concordant and statistically significant associations with all three menopause factors (Fig. 3c). We performed GO and pathway enrichment to elucidate whether there may be distinct biological processes associated with proteins shared among menopause timing and menopause type and/or stage, compared to proteins unique to menopause timing. Of the proteins unique to menopause timing, those positively associated with age at menopause (*N*=67 proteins) were enriched for processes involving lipid metabolism, synaptic signaling, and cellular adhesion/growth signaling, while those negatively associated with age at menopause (*N*=201 proteins) primarily reflected inflammatory biology (Fig. 3d). By contrast, of the proteins shared between menopause timing and type/stage, those positively associated with age at menopause were enriched for hormone-responsive extracellular signalling and developmental biology (*N*=18 proteins), while those negatively associated with age at menopause were linked to cell surface and plasma membrane immune processes (*N*=378 proteins; Fig. 3e). Collectively, these findings suggest that despite some shared convergence on core reproductive and endocrine biology, the proteomic signatures of menopause timing are dissociable from those of surgical and spontaneous menopause.

### Sensitivity analyses

Sensitivity analyses in the UKB showed that the proteomic correlates of menopause timing were robust to hormone therapy use, multiple chronic health conditions and comorbidities, key demographic factors, *APOE4* status, and restriction of the sample to women aged ≥65 years (Extended Data).

### Earlier menopause proteins predict future dementia risk and poorer brain health outcomes in postmenopausal women

We next evaluated whether proteins associated with menopause timing in the UKB associated with incident all-cause dementia in the same participants over a mean (*SD*) of 15.7 (2.95) years of follow-up (*N*=546 dementia cases; Fig. 4a). Confirming previous findings in the UKB,^21^ age at menopause was associated with risk for incident dementia (HR=0.97, *p*<.001), such that women with the earliest tertile of age at menopause had a hazard ratio of 1.37 for progressing to dementia relative to the latest tertile. Of the 668 menopause timing proteins, 175 proteins associated with greater risk of incident dementia and 28 proteins associated with lower risk of dementia in age-adjusted Cox proportional-hazards models (Fig. 4b; Tab.S17). Menopause timing proteomic effect sizes exhibited a strong negative correlation with proteomic effect sizes for incident dementia (r=-0.40, Fig. 4c) such that proteins associated with later menopause were generally associated with lower risk of dementia and vice-versa.

**Figure 4.**
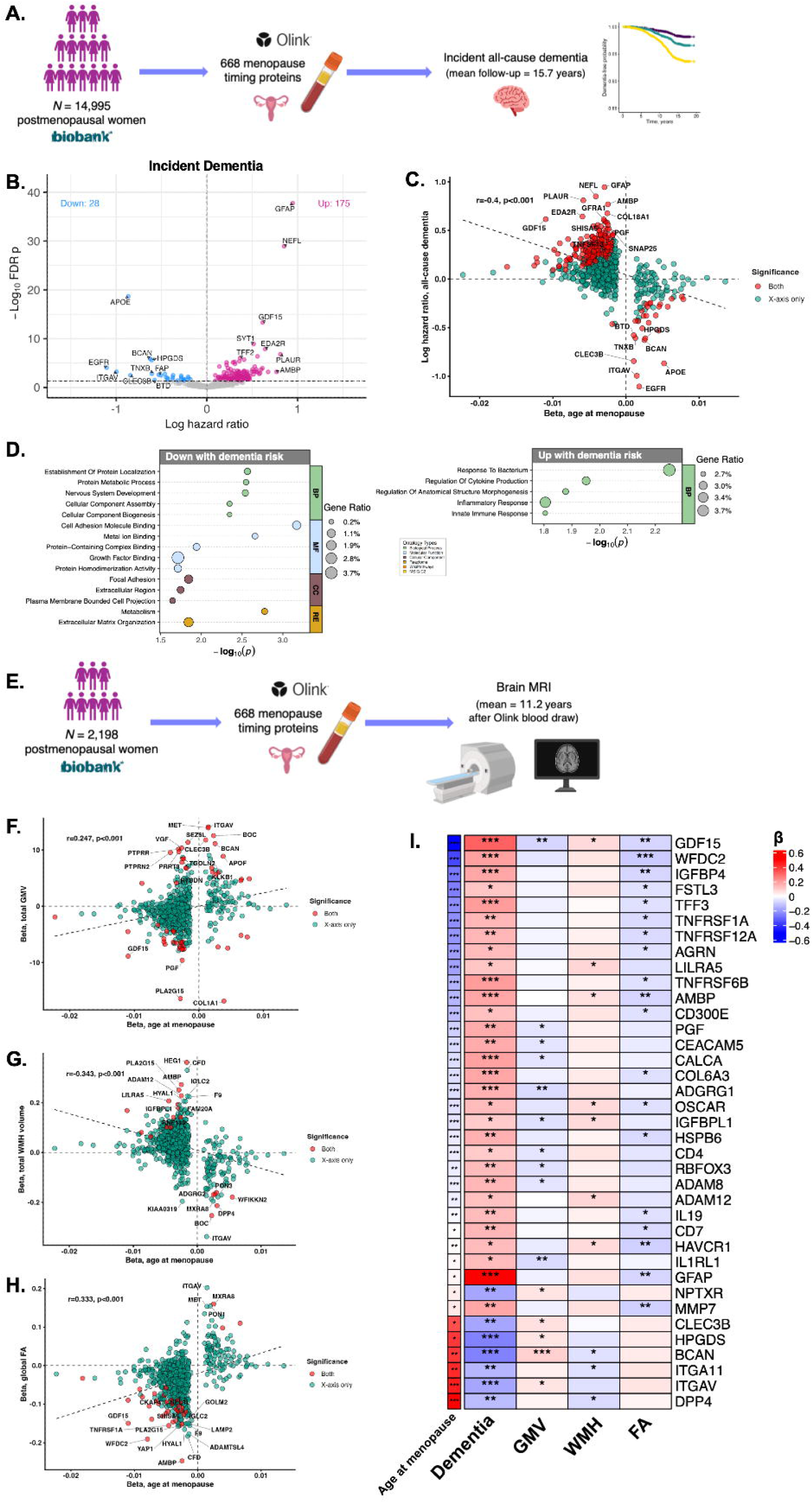
Menopause timing proteins overlap with those implicated in dementia risk and brain aging. **A. Overview for dementia analyses.** Age-adjusted Cox proportional hazards models tested associations of menopause timing proteins (*N*=668) with incident all-cause dementia over a mean of 15.7 years of follow-up. **B. Protein associations with incident dementia.** Of the 668 menopause proteins, 175 were associated with increased dementia risk, and 28 with decreased dementia risk (FDR *p*<.05). **C. Comparison of protein effect sizes for menopause timing and dementia risk.** There was strong negative concordance between menopause timing proteomic shifts and those associated with dementia. Proteins that were positively associated with age at menopause were associated with lower risk of incident dementia, and proteins that were negatively associated with age at menopause were associated with higher risk of incident dementia. **D. Gene ontology (GO) and pathway enrichment for menopause timing proteins that also associate with dementia risk.** Menopause proteins may associate with dementia risk via upregulation of inflammatory processes and downregulation of adaptive cholesterol regulation and extracellular matrix maintenance. **E. Overview for brain magnetic resonance imaging (MRI) analyses.** *N*=2,198 postmenopausal women in our analytic sample underwent brain MRI at a mean (*SD*) of 11.22 (3.06) years after the Olink proteomics blood draw. Linear models tested associations between menopause timing proteins (*N*=668) and brain outcomes, adjusted for age at baseline and time between Olink blood draw and MRI scan. **F-H. Comparison of protein effect sizes for menopause timing and MRI brain outcomes.** There was concordance between menopause timing proteomic shifts and those associated with all three brain outcomes. Proteins that were positively associated with age at menopause were associated with larger GMV, lower WMH burden, and higher FA, and proteins that were negatively associated with age at menopause were associated with smaller GMV, greater WMH burden, and lower FA. **I. Heatmap of menopause proteins that most consistently associate with dementia and brain outcomes.** Of the menopause timing proteins that were significantly associated with dementia risk (*N*=668), 37 were also associated with at least one MRI brain outcome. The heatmap shows effect sizes (standardized betas for cross-outcome comparison) and statistical significance (FDR *p*) for these 37 proteins’ associations with all four cognitive/brain outcomes, annotated by age at menopause effect size.

Higher GDF15, previously linked to dementia risk in sex-aggregated UKB studies,^41,42^ was associated with greater risk of dementia such that relative to those in the lowest tertile, women with the highest tertile of GDF15 had a hazard ratio of 1.70 for progressing to dementia. GFAP and NEFL, established neurodegenerative markers, exhibited more modest but statistically significant increases with earlier menopause and were the strongest predictors of greater dementia risk. By contrast, higher APOE was associated with later menopause and the strongest predictor of lower dementia risk. GO and pathway analysis for overlapping menopause timing and dementia risk proteins (Fig. 4d) showed enrichment of immune-related proteins in earlier menopause/higher dementia risk (e.g., GDF15, TNFSF13B, PLAUR) and enrichment of proteins involved in lipid metabolism (e.g., APOE, HPGDS, PON1) and nervous system development in later menopause/lower dementia risk (e.g., BCAN, LEFTY2).

To contextualize findings, we also evaluated whether menopause timing proteins showed sex-specific associations with incident all-cause dementia in all UKB women and men ≥50 years old (*N*=34,325; *N*[%] men=15,776[46.0], mean(*SD*) age=60.3[5.50]). Of the 668 menopause timing proteins, 48 showed FDR significant interactions with sex on dementia risk in age-adjusted Cox models (Tab. S18; Fig. S9a). Simple slopes analysis revealed that 44 of these 48 proteins were significantly positively associated with dementia risk in women and not men (Fig.S9b-e), including serine protease CFD, cytokine CYTL1, and PLAUR. Of note, CFD is a key activator of the alternative complement pathways and is highly abundant in fat tissue.^43^ Additionally, GDF15 and GFAP showed stronger positive associations with dementia risk in women relative to men.

In a subset of postmenopausal women with brain MRI (*N*=2,198), age-adjusted linear models examined whether the 668 menopause timing proteins associated with MRI-derived measures reflecting neurodegeneration, cerebral small vessel disease burden, and white matter microstructural integrity (Fig. 4e). There were 25 proteins positively associated with total grey matter volume (GMV) and 28 negatively associated with total GMV; 14 proteins positively associated with white matter hyperintensity (WMH) burden and 6 proteins negatively associated with WMH; and 3 proteins positively associated with global fractional anisotropy (FA) and 35 proteins negatively associated with global FA (Tabs.S19-21, Figs.S10-12). Menopause timing proteomic effect sizes showed concordance with brain MRI effect sizes across all three outcomes (GMV: r=0.247, WMH: r=-0.343, FA: r=0.333). Specifically, proteins that were positively associated with age at menopause were associated with larger GMV, lower WMH burden, and higher FA, and proteins that were negatively associated with age at menopause were associated with smaller GMV, greater WMH burden, and lower FA (Figs.4f-h). GDF15 and BCAN, top menopause timing targets associated with dementia risk, exhibited some of the strongest relationships across multiple neuroimaging measures (Fig. 4i).

## DISCUSSION

We comprehensively mapped molecular signatures of menopause timing in blood and identified their overlap with later-life brain aging and dementia risk. We discovered widespread and replicable proteomic shifts linked to earlier menopause, reflecting pro-inflammatory, extracellular matrix remodeling, and apoptotic pathways. We showed that earlier menopause is associated with accelerated biological aging across proteomic clocks of multiple organs and cell types, with brain aging among the top effect sizes. We additionally observed meaningful overlap between proteins associated with menopause timing and those implicated in brain aging, including inflammatory markers associated with earlier menopause and greater dementia risk (e.g., GDF15), as well as neuroplasticity factors associated with later menopause and cognitive preservation (e.g., BCAN). Collectively, our findings suggest that age at menopause may be a meaningful biomarker of women’s biologic age. We established a proteomic signature of menopause timing that reflects potential molecular drivers of increased risk for age-related diseases in women with earlier menopause, highlighting this midlife transition as a critical inflection point for later-life brain health trajectories.

Across both cohorts, GDF15 was among the top targets upregulated with earlier menopause, an association that persisted after adjustment for demographic factors and multiple age-related diseases and cardiometabolic conditions. Elevated GDF15 in blood strongly prognosticates increased risk for a wide range of age-related diseases, including dementia.^42,44^ Accordingly, GDF15 has recently been proposed as a proxy biomarker for epigenetic aging.^45^ Very limited research has examined GDF15 in the context of menopause. Our recent data show higher plasma GDF15 levels in postmenopausal women relative to age-matched premenopausal women,^20^ consistent with previous observations of accelerations in biological aging across the menopause transition.^28,46^ Our study further extends these findings to suggest GDF15 may play a role in signaling and/or be a downstream readout of menopause timing itself. Beyond GDF15, the proteomic profiles of earlier menopause were largely distinct from those of menopause type and stage. These findings support the presence of distinct coordinated biological systems across menopause features and highlight early menopause specifically as a signal of elevated susceptibility to age-related disease processes.

Brain aging signals were consistently represented among the menopause timing proteome, including overlap with targets associated with incident dementia and poorer brain structure. Across multiple outcomes in the UKB, proteins upregulated in earlier menopause associated with higher risk for dementia and worse MRI-derived measures of brain health, while the reverse patterns held for proteins upregulated in later menopause. Furthermore, of the menopause timing proteins that showed sex-specific associations with dementia risk, the majority (44/48 proteins) predicted greater risk in women than men, supporting the hypothesis that female-specific endocrine processes may contribute to sex differences in dementia outcomes.^4,16–20^ Together, these findings align with accumulating data pinpointing earlier menopause as a key dementia risk factor,^5,6,21,22^ shedding light on molecular pathways that may link menopause timing to neurodegenerative risk. Several top targets included the inflammatory molecules GDF15, CHI3L1 (YKL-40), and PLAUR. We also observed statistically significant associations for both earlier menopause and dementia risk with established AD biomarkers GFAP, NEFL, and SMOC1, though the effect sizes for menopause timing were relatively small. By contrast, top targets linked to both later age at menopause and lower dementia/neurodegenerative risk included the cholesterol transporter APOE and neurodevelopmental and neuroplasticity factors WFIKKN2 and BCAN. Later menopause may confer protection against neurodegenerative processes partly via synaptic resilience,^6,47,48^ possibly mediated by longer exposure to estradiol and/or other more youthful neurohormonal profiles (e.g., lower FSH).^49^

It is unclear whether earlier menopause causally induces faster rates of biological and brain aging, perhaps via earlier changes to the hormonal and ovarian signalling milieu,^50^ or whether earlier spontaneous menopause is a consequence of a broader biological system primed for accelerated aging, potentially coordinated via upstream mechanisms such as genetically determined intrinsic aging pace.^51^ There is extensive evidence from preclinical models that surgical or medical induction of menopause results in widespread detrimental health effects.^52–58^ In humans, bilateral oophorectomy is similarly a significant risk factor for multisystem morbidity including dementia.^59–61^ Together, these results implicate a causal role for earlier menopause timing in inducing accelerated aging trajectories. However, it is also possible that earlier spontaneous menopause also reflects a broader coordinated system of accelerated endogenous aging pace. Emerging data suggest hypothalamic aging may precede ovarian senescence and trigger menopause,^12,13^ pointing to upstream and possibly brain-mediated factors regulating menopause timing. Indeed, among our aging clock findings, we observed some of the strongest associations between menopause timing proteomics and brain aging measures. Menopause timing biology may therefore serve as a discovery framework for identifying intervention targets for age-related diseases, including dementia.

Strengths of this study include the use of large cohorts with plasma proteomics to define replicable molecular signatures of menopause timing and their associations to brain aging outcomes. There were also limitations, such as self-report data on menopause history, which may introduce imprecision and/or recall bias. Earlier age at menopause is associated with increased mortality risk,^26^ which may introduce survival bias. Limited data on hormone therapy in the UKB precluded a more detailed investigation of the effects of hormone therapy on menopause-related proteomic shifts. Finally, while the small but statistically significant associations of earlier menopause with elevations in established neurodegenerative biomarkers (e.g., GFAP, NEFL) are intriguing, we did not have orthogonal AD biomarkers to further test these associations.

We identified a replicable proteomic signature of menopause timing reflecting accelerated multi-system aging and identified oligodendrocyte, inflammatory, lipid, and neurotrophic (e.g., ECM, growth factor) pathways as possible links connecting menopause timing to brain aging and dementia risk. These findings underscore the ability to leverage the menopause transition as a discovery tool to disentangle fundamentals of aging biology and highlight menopause timing biology as a potential target to reduce risk of dementia.

## METHODS

### Discovery Cohort: UKB

Our discovery cohort consisted of *N*=15,012 postmenopausal women in the UKB with proximity extension assay Olink plasma proteomics data for up to 2,923 proteins (Tab.S1).^62,63^ Cohort details are described in the Extended Data. To mitigate associations between chronological age and age at menopause introduced by sampling bias, we restricted the sample to women ≥50 years old, resulting in a minimal correlation between chronological age and age at menopause (r=0.079). Incident all-cause dementia was defined algorithmically,^34^ and participants were censored at first dementia diagnosis, death, or September 2025. A subset (*N*=2,198) underwent 3T brain MRI at a mean (*SD*) of 11.22 (3.06) years after the Olink blood draw. We considered three global measures (Tab.S22) reflecting brain aging and neurodegenerative risk: (1) GMV (normalized for head size); (2) total WMH volume, scaled for head size and log-transformed;^64^ and (3) global FA (Extended Data).^65^

### Validation Cohort: WHI-LLS

To validate menopause timing findings, we used data from *N*=1,210 postmenopausal women in the WHI-LLS^66,67,68^ who had Olink Target plasma proteomics for 540 unique proteins, 424 of which overlapped with those available in UKB (more details in Extended Data).^69^ Of these participants, *N*=470 (38.8%) were randomized to hormone therapy in the original WHI clinical trial, *N*=470 (38.8%), *N*=440 (36.4%) were randomized to placebo, and *N*=300 (24.8%) were in the WHI observational study only (Tab.S3). The correlation between age at menopause and chronological age in this sample was similar to UKB (r=0.087).

All cohorts were approved by research ethics boards and all participants provided informed consent.

### Analyses

For all main analyses, models were adjusted for chronological age, and *p*-values controlled for FDR at 5% using the Benjamini-Hochberg method. GO and pathway enrichment were performed using the publicly-available GOparallel code (<u>github.com/edammer/GOparallel</u>) where specified.^70^ Effect size correlation plots were used to examine concordance across summary statistics where specified.

Across both the UKB and WHI-LLS, linear models tested the associations of age at menopause with protein levels, adjusting for chronological age and original WHI randomization status (i.e., active treatment vs. placebo/observational study only; in WHI-LLS only). To assess the links between menopause timing with biological aging, in the UKB, we calculated proteomic aging clocks for 13 organ systems and 38 cell types using previously established Olink models (Extended Data).^34,35^ Because the organ age models did not include a measure of reproductive aging, to evaluate whether menopause timing proteins may originate disproportionately from female reproductive tissue, we used Fisher’s exact test to evaluate whether proteins that may have originated from female reproductive tissue (per Human Protein Atlas classifications; Extended Data)^71^ were overrepresented among targets that associated with age at menopause.

While menopause timing may generally influence brain aging, surgical menopause is associated with especially pronounced increases in dementia risk.^7,60^ To address this, in the UKB, we tested associations of menopause type (surgical/spontaneous) with proteins, adjusting for age at menopause, hormone therapy history, and age. To further disentangle the proteomic correlates of menopause timing from those of the menopause transition itself, we further compared the age at menopause–protein association effect sizes to summary statistics describing differential protein abundance in age-matched pre- and postmenopausal women with no history of surgical menopause or current hormone therapy in the UKB (*N*=2,814).^20^

To assess the relevance of menopause timing-related proteomic shifts to dementia risk, in the UKB, we tested the associations of menopause proteins with incident all-cause dementia using age-adjusted Cox proportional-hazards models. We also tested associations of menopause proteins with MRI-derived measures of total GMV, WMH burden, and global FA, using parallel linear models adjusted for age at Olink blood draw and time between blood draw and MRI visit.

## Data Availability

All data produced in the present study are available to qualified investigators upon request to the UK Biobank and Women’s Health Initiative.

## Funding/Acknowledgments

This work was supported by the Wellcome Leap CARE program (MPI: KBC, RS, EGJ), RF1AG096165 (KBC); the Ann S. Bowers Women’s Brain Health Initiative (KBC, EGJ); R01AG072475 (PI: KBC), and K23AG090757 (PI: RS). Our work is also supported by a grant from the Larry L. Hillblom Foundation (2024-A-001-CTR; PI: KBC), New Vision Research (CCAD 2024-001-1; PI: RS), an award from the American Academy of Neurology (PI: RS), funding from the Alzheimer Society of Canada Research Program (MWA, JSR), CIHR (173253, 438475, Canada Graduate Scholarships program; JSR, MWA), the Alzheimer’s Association (JSR). This work was supported in part by NIA Intramural Research Program (K.A.W.). The contributions of the NIH author(s) are considered Works of the United States Government. The findings and conclusions presented in this paper are those of the author(s) and do not necessarily reflect the views of the NIH or the U.S. Department of Health and Human Services. This research has been conducted using the UK Biobank Resource under Application Number 1045037. The WHI program is funded by the National Heart, Lung, and Blood Institute, National Institutes of Health, U.S. Department of Health and Human Services through contracts 75N92021D00001, 75N92021D00002, 75N92021D00003, 75N92021D00004, 75N92021D00005. Drs. Kooperberg and Reiner, and the WHI-LLS proteomic measurements, were supported by grant R01 HL136574. Work on *The effects of pharmacological and spontaneous menopause on Alzheimer’s risk in women* is supported by Wellcome Leap as part of the CARE Program. Plots were created in part using Biorender.

## SHORT LIST OF WHI INVESTIGATORS

Program Office: (National Heart, Lung, and Blood Institute, Bethesda, Maryland) Jared Reis and Candice Price Clinical Coordinating Center: (Fred Hutchinson Cancer Center, Seattle, WA) Garnet Anderson, Charles Kooperberg, and Holly Harris Steering Committee: (Fred Hutchinson Cancer Center) Marian Neuhouser – Committee Chair; (Fred Hutchinson Cancer Center) Garnet Anderson; (University of California, Davis) Lorena Garcia; (Wake Forest University) Lindsay Reynolds; (University at Buffalo) Amy Millen; (University at Buffalo) Jean Wactawski-Wende; (Fred Hutchinson Cancer Center) Holly Harris; (University of Massachusetts) Brian Silver; (University of Tennessee Health Center) Karen Johnson; (Stanford Prevention Research Center) Marcia L. Stefanick; (The Ohio State University) Electra Paskett; (Wake Forest University School of Medicine) Mara Vitolins

