## Extended data for "Plasma proteomics link menopause timing to brain aging and dementia risk"

**Sensitivity analyses show that the proteomic correlates of menopause timing were robust to hormone therapy use, chronic health conditions, and key demographic factors.**

To assess the potential role of hormone therapy use, which was more common in women with earlier menopause (Table S1), we compared effect sizes of age at menopause from our primary, age-adjusted models to those of age at menopause from our multivariable models adjusted for age, menopause type (spontaneous/surgical), and hormone therapy history (i.e., current user, past user, never used). Effect sizes demonstrated very strong concordance, suggesting that hormone therapy use does not confound the observed associations between age at menopause and the proteome (Table S11, Fig.S4). We further evaluated the associations of age at menopause with protein levels in models stratified by history of hormone therapy use (i.e., ever used, *N*=6,317; never used, *N*=8,059), adjusting for age and type of menopause. Effect sizes again demonstrated strong concordance, suggesting similar effects of menopause timing on the plasma proteome between individuals with and without histories of hormone therapy use (Tables S12 & S13, Fig.S5).

In additional sensitivity analyses, we re-evaluated associations of age at menopause with protein levels additionally adjusting models for a range of clinical and health history variables that may influence dementia risk (i.e., body mass index (BMI), low density lipoprotein (LDL) cholesterol levels, smoking status, systolic and diastolic blood pressure, hbA1C, plus histories of stroke, angina, heart attack, broken bones within the past 5 years, and cancer; *N*=12,533). These analyses yielded very similar results to the main models (Table S14, Fig.S6), suggesting that prevalent age-related health conditions do not account for the proteomic shifts associated with menopause timing. In a second set of models, we re-estimated associations of age at menopause with protein levels additionally adjusting for *APOE4* carriage and years of education, which also produced highly consistent results (*N*=10,099, Table S15, Fig.S7). Finally, to ensure the proteomic correlates of menopause timing were independent of residual confounding by chronological age, we tested associations of age at menopause with protein levels in a restricted subsample of women aged ≥65 years, which also yielded very similar results (*N*=3,885, Table S16, Fig.S8).

**Discovery Cohort Details: UK Biobank**

The UK Biobank is a population-based prospective cohort study with clinical data on approximately 500,000 participants aged 40-69 years at baseline.^1^ The UKB-PPP consortium generated proximity extension assay (PEA) Olink plasma proteomics data on a subset of participants’ baseline samples, resulting in up to 2,923 protein measurements from 52,995 participants in the post-UKB-PPP quality control dataset.^2^ Details of proteomics data generation, processing, and quality control have been previously described^2^ and are available at <https://biobank.ndph.ox.ac.uk/ukb/ukb/docs/PPP_Phase_1_QC_dataset_companion_doc.pdf>. We excluded 9 proteins with >20% missing data, for a total of 2,914 proteins in main analyses.

Of participants with baseline Olink proteomics data, there were *N*=16,286 female participants who reported being postmenopausal at baseline (answered “yes” to "Have you had your menopause (periods stopped)?") and had available data on self-reported age menopause, which in the UKB is defined as age at final menstrual period. Of postmenopausal women with age at menopause data, we excluded *N*=533 postmenopausal women who were younger than 50 years old at baseline to mitigate associations between chronological age and age at menopause introduced by sampling bias, resulting in a minimal correlation between chronological age and age at menopause (r=0.079). To ensure that age at menstrual cessation accurately reflects age at ovarian failure, we further excluded women who reported having a hysterectomy at a younger age than their age at final menstrual period, with either no bilateral oophorectomy (*N*=696) or bilateral oophorectomy at an age after age at hysterectomy (*N*=45). These exclusions resulted in a total main analytic sample of *N*=15,012 (Table S1). Surgical menopause was defined as bilateral oophorectomy at an age equal to or younger than self-reported age at final menstrual period, spontaneous (i.e., natural) menopause was defined as menstrual cessation in women with no history of bilateral oophorectomy. In addition to age at menopause and cause of menopause, participants self-reported their histories of hormone therapy use, demographic information, and medical conditions. Proteomic analyses for menopause type included *N*=14,772 women with available data on menopause type and hormone therapy history (*N*=208 missing menopause type, *N*=32 missing hormone therapy data).

Consistent with prior work, incident all-cause dementia was algorithmically defined using a combination of information in international classification of diseases (ICD) diagnosis codes in linked primary care, hospital inpatient, and death registry data (codes A81, F00 to F03, F05, F10, G30, G31 and I67).^3^ Analyses of incident dementia included a subset of *N*=14,995 participants (*N*=17 with dementia at baseline excluded). Participants were censored at first dementia diagnosis, death, or September 2025, representing the most recent update of algorithmically defined dementia outcomes in the UKB.

A subset of participants in our main analytic sample (*N*=2,198) underwent brain MRI on a standard Siemens Skyra 3T scanner with a 32-channel head coil, including T1w and T2w structural imaging and diffusion imaging. Brain MRI was acquired at a mean (*SD*) of 11.22 (3.06) years after the Olink blood draw. Image acquisition parameters and processing pipelines have been previously described^4^ and are presented in detail at [biobank.ndph.ox.ac.uk/showcase/showcase/docs/brain_mri.pdf](https://biobank.ndph.ox.ac.uk/showcase/showcase/docs/brain_mri.pdf). In brief, MRI quality control and analysis is performed using an automated pipeline at the University of Oxford’s Wellcome Centre for Integrative Neuroimaging, which primarily calls FSL and FreeSurfer tools.^5^ We considered three global measures reflecting brain aging and neurodegenerative risk: (1) total grey matter volume (normalized for head size); (2) total volume of white matter hyperintensities (WMH) from T1w and T2w fluid-attenuation inversion recovery (FLAIR) images, scaled for head size and log-transformed;^6^ and (3) global fractional anisotropy (FA), computed as the average of standardized mean diffusion MRI FA values from 48 white matter tracts (Table S22).^7^

**Validation Cohort Details: Women’s Health Initiative (WHI) Long-Life Study (LLS)**

To validate menopause timing findings, we used data from the WHI LLS Visit 1 (2012-13). The LLS is an extension of the original WHI study^8,9^ that recruited a subset of *N*=7,875 surviving women aged 63 to 99 years for clinical evaluation and blood collection.^10^ Of these participants, *N*=1,388 had Olink Target plasma proteomics from six panels (Cardiometabolic, Cardiovascular II, Cardiovascular III, Inflammation, Neurology, and Oncology III), resulting in 540 unique protein measurements, 424 of which overlapped with those available in UKB.^11^ Protein levels were winsorized at 5 times the standard deviation (*SD*) above or below the mean. All women were postmenopausal at the time of original WHI enrollment (1993-98), and female reproductive health history factors were self-reported. In the WHI, age at menopause is determined using self-report data following an established algorithm. In brief, age at menopause was defined as the age at which she first experienced any of the following: (1) last had any menstrual bleeding; (2) had a bilateral oophorectomy; or (3) began using hormone replacement therapy. If a woman had hysterectomy without bilateral oophorectomy, her age at menopause was defined as either (1) the age she began using hormone replacement therapy; or (2) first had symptoms of menopause, such as hot flashes or night sweats. Further details on age at menopause ascertainment are available at: whi.org/doc/Algorithm-Age-at-Menopause.pdf.

Our analytic sample included *N*=1,210 participants with proteomics and available age at menopause (Table S3; *N*=126 with unknown age at menopause excluded). The correlation between age at menopause and chronological age in this sample was similar to UKB (r=0.087).

**Organ aging and cell aging models**

To assess the links between menopause timing with biological aging, in the UKB, we calculated proteomic aging clocks for 13 organ systems and 38 cell types using previously established Olink models.^3,12^ Organ and cell clock models were developed using proteins that are labeled as enriched for the corresponding tissue or cellular sources in the Human Protein Atlas (HPA),^13^ such that proteins that load onto each organ- or cell-specific model are assumed to originate at least in part from those organs/cells.^3^ Because the organ and cell clock estimations required complete data, these analyses were performed on a subset of *N*=12,716 postmenopausal women following K nearest-neighbours imputation of the Olink data (below).^3^ Chronological age-adjusted linear models tested associations of age at menopause with organ and cell age “gaps” (i.e., z-scored residuals of proteomic predicted age linearly regressed against actual age, whereby larger gaps reflect more advanced biological aging).

**Female tissue enrichment**

Because the organ age models did not include a measure of reproductive aging, to evaluate whether menopause timing proteins may originate disproportionately from female reproductive tissue, we used HPA classifications to create a supraordinate “female reproductive tissue” category.^13^ We included proteins that were classified “tissue-enriched” or “group-enriched” (whereby RNA-seq number of transcripts per million (nTPM) ≥4 times higher in the enriched tissues vs. remaining tissues) for the endometrium, ovary, fallopian tube, cervix, vagina, breast, or placenta. Then, we used Fisher’s exact test to evaluate whether proteins that may have originated from female reproductive tissue were overrepresented among targets that either positively or negatively associated with age at menopause.

**UK Biobank Olink imputation for organ age models**

We imputed Olink proteomics data using the entire Olink baseline sample (*N*=52,995 participants; *N*=2,923 proteins) following a previously published method.^3^ For quality control, we first excluded *N*=8,181 samples with more than 1,000 proteins missing, seven proteins with missing values in more than 10% of samples, and 338 samples with discordant self-reported sex and genetic sex, resulting in a final dataset of *N*=44,476 participants with 2,916 protein measurements. We performed missing value imputation of the Olink data using *k*-nearest neighbors (KNN) imputation. We split the data into train and test, where each split comprised 11 randomly selected UKB assessment centers (train centers: Newcastle, Nottingham, Bristol, Leeds, Sheffield, Glasgow, Wrexham, Croydon, Hounslow, Edinburgh; test centers: Liverpool, Manchester, Middlesborough, Oxford, Stockport, Stoke, Swansea). Protein levels were z-transformed using the means and standard deviations of the train split. Using scikit-learn’s KNNImputer function (scikit-learn.org), we trained a KNN imputed where the number of neighbours (*k*) was set to the square root of the sample size in the train split (*k* = 166). We assessed the imputer’s performance on *N*=5,590 samples (3,425 train/2,165 test) with no original missing protein values. To do so, we randomly input missing values into this “ground truth” subsample at 3% (i.e., equivalent to the missing value rate in the entire post quality control dataset). We then used the imputer on this subsample to calculate the error between imputed values and ground truth values. This resulted in a total mean error rate (MAE) of 0.57 in both the train and test samples, consistent with previous research^3^ and supporting robust imputation of the Olink data (Fig.S13).

**Figure S1. Comparison of effect sizes of age at menopause on protein levels in main, age-adjusted WHI-LLS models (x-axis) and in models additionally adjusted for race and ethnicity.**

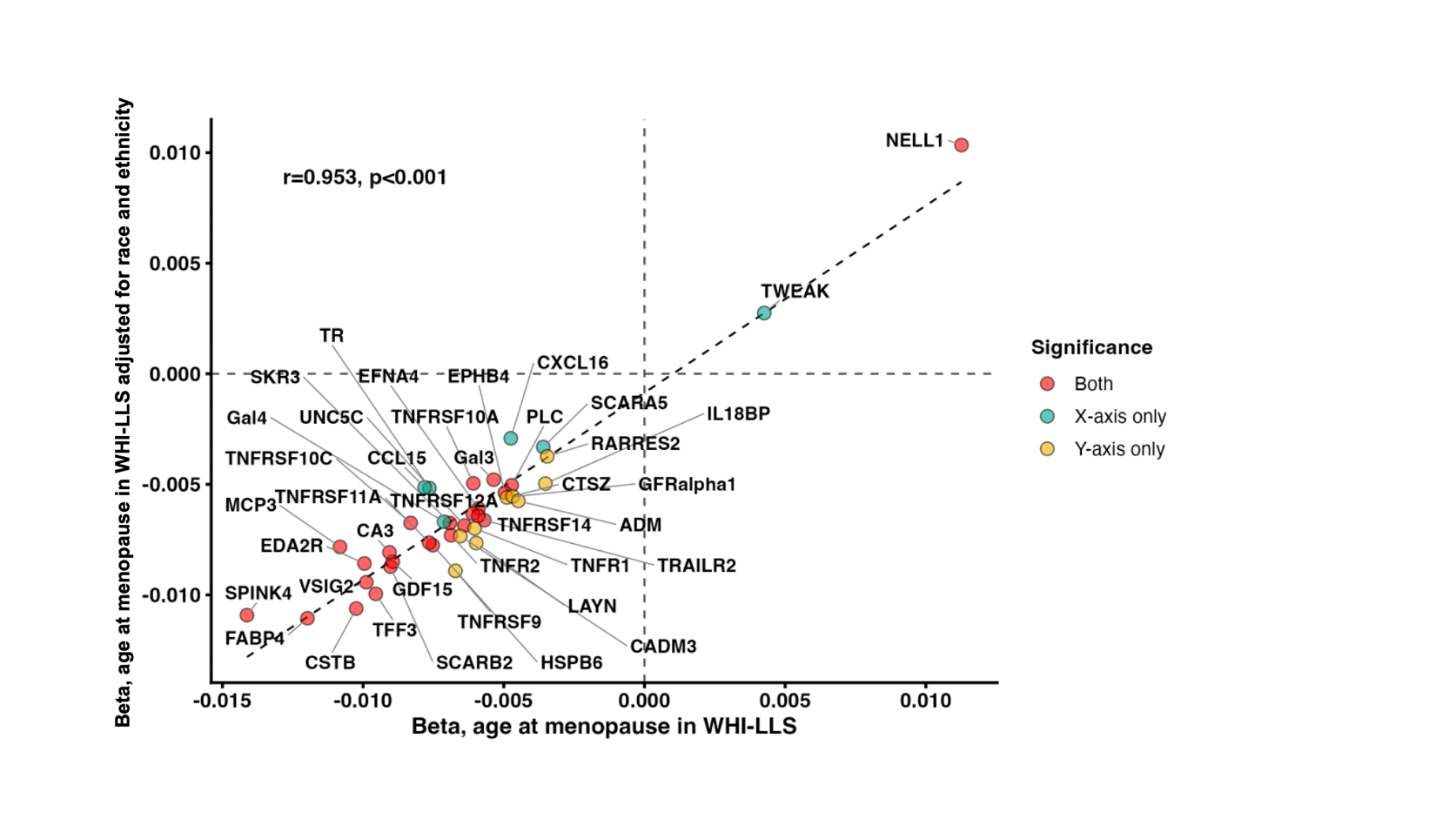

**Figure S2. Cell age in UK Biobank (UKB). Linear models adjusted for age showed that age at menopause was associated with smaller (i.e., more favourable) age gaps on 35 of the 38 cellular aging measures (FDR *p*<.05).**

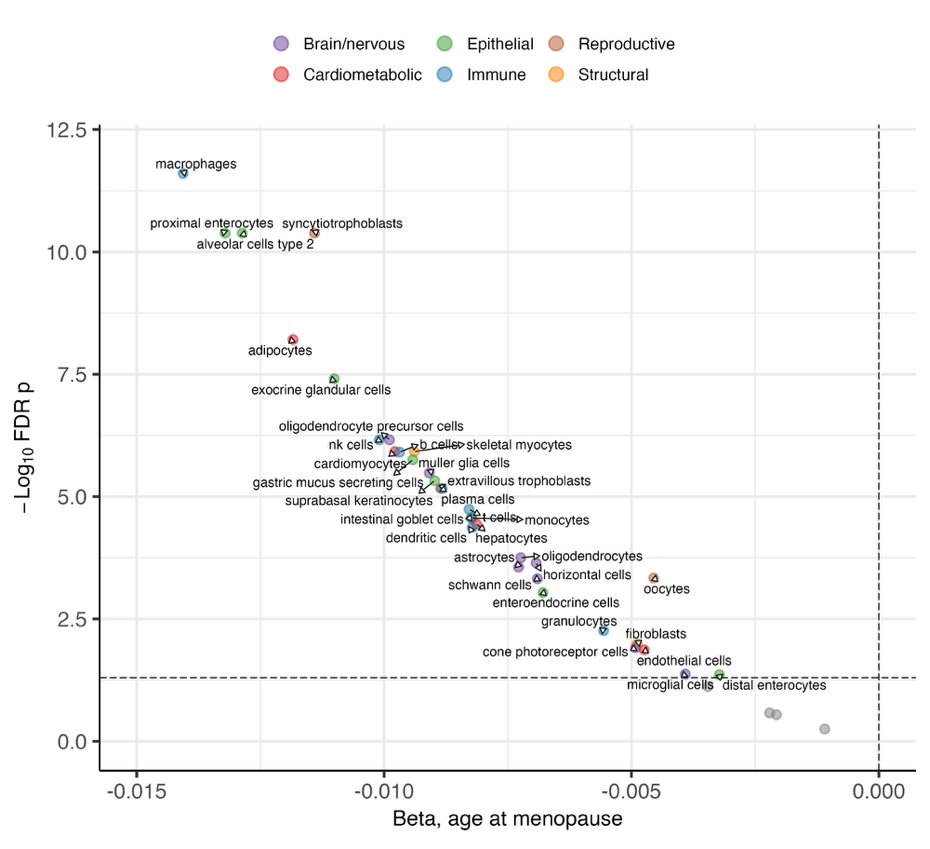

**Figure S3. Differential abundance by cause of menopause. Multivariate linear models adjusted for age, age at menopause, and history of hormone therapy showed that surgical menopause was associated with upregulation of 55 proteins and downregulation of 33 proteins (FDR *p*<.05).**

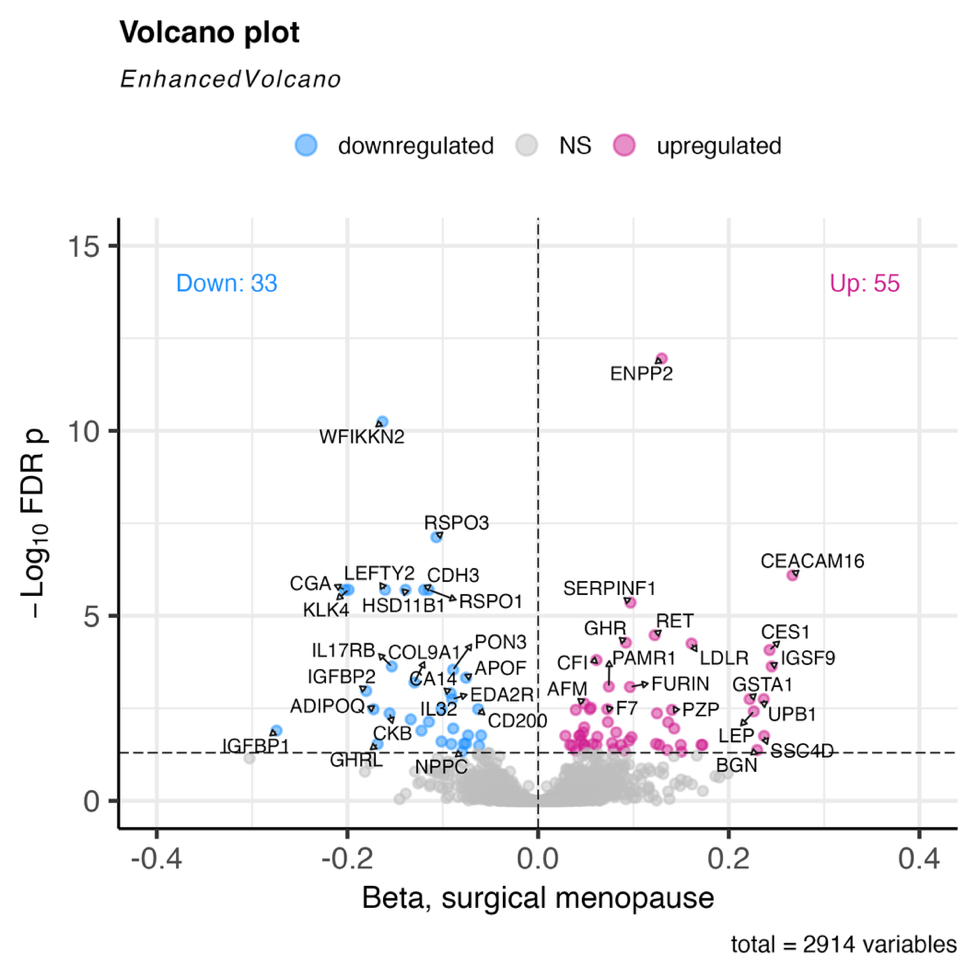

**Figure S4. Comparison of effect sizes of age at menopause on protein levels in main, age-adjusted models (x-axis) and in models additionally adjusted for cause of menopause and history of hormone therapy (HT) use (y-axis).**

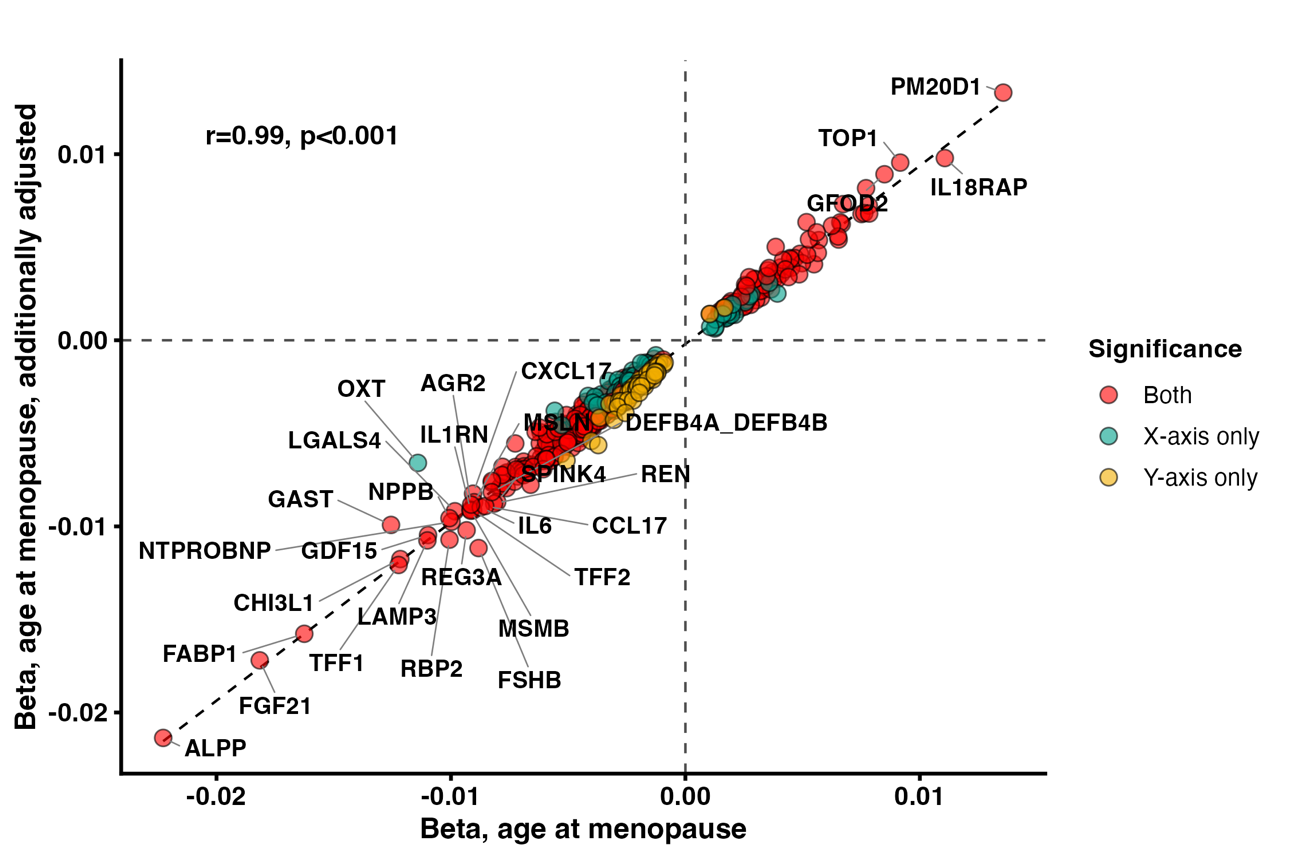

**Figure S5. Comparison of effect sizes of age at menopause on protein levels stratified by women with no history of hormone therapy (HT; x-axis) and women with history of HT use (y-axis).**

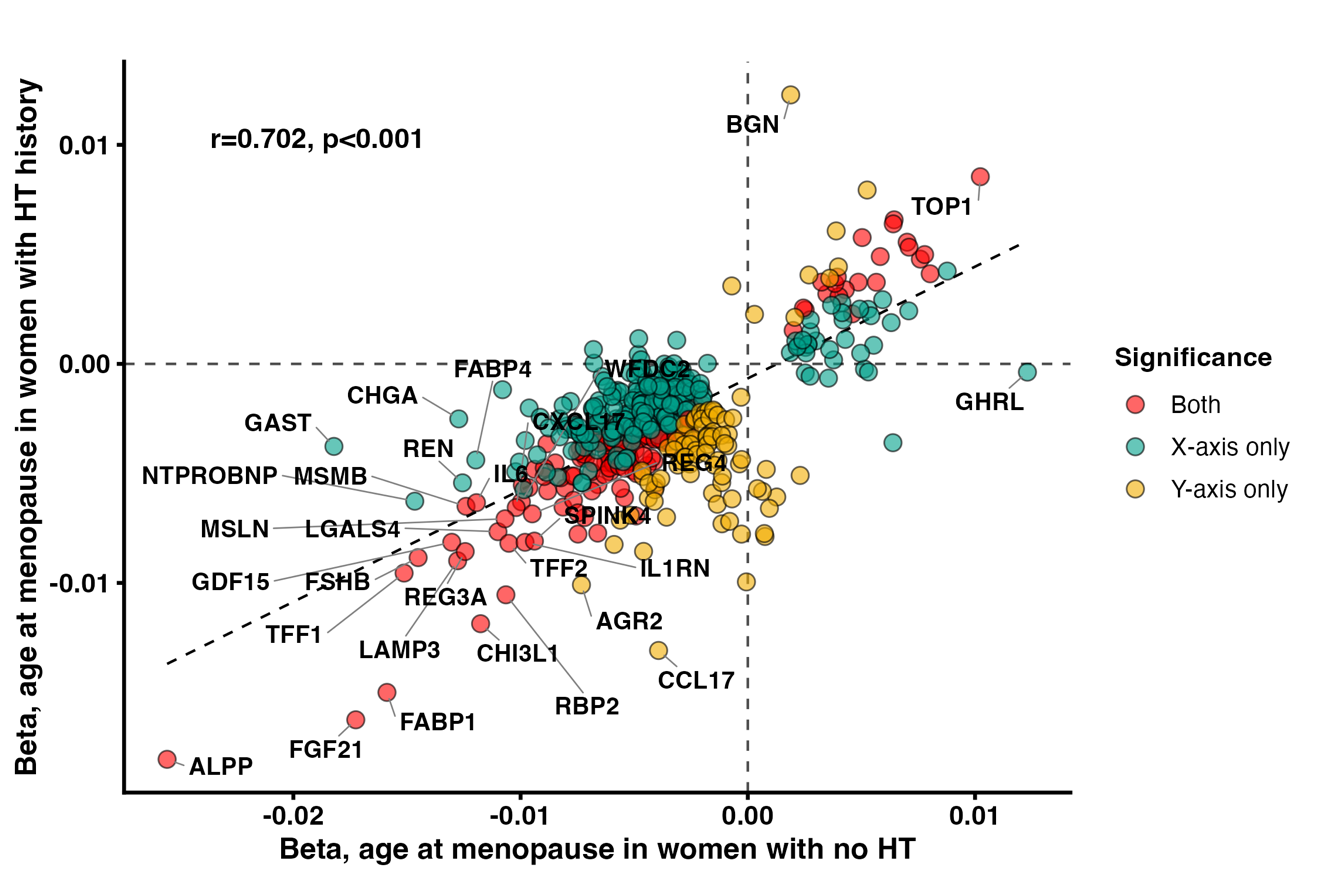

**Figure S6. Comparison of effect sizes of age at menopause on protein levels in main, age-adjusted models (x-axis) and in models additionally adjusted for cardiometabolic risk factors and chronic conditions (y-axis).**

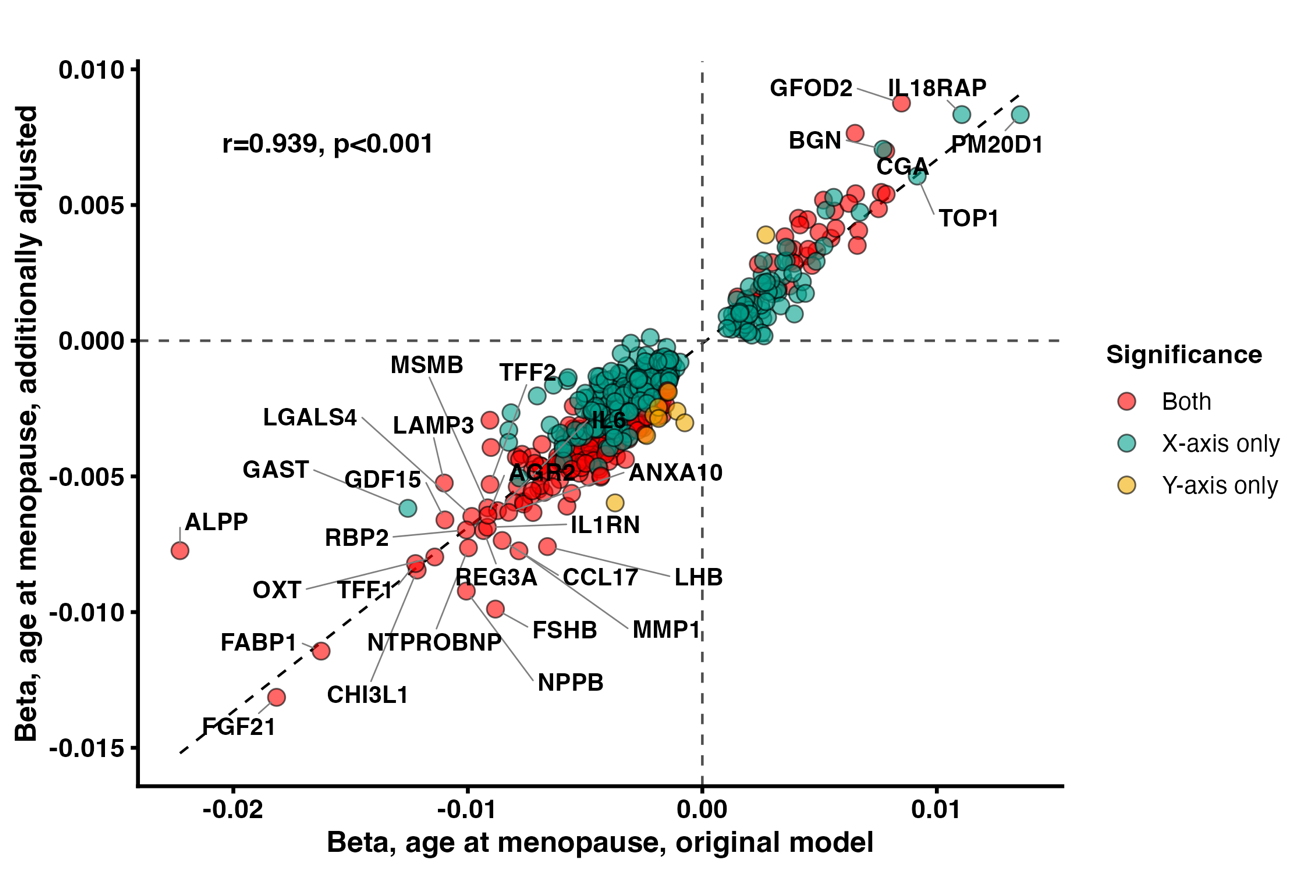

**Figure S7. Comparison of effect sizes of age at menopause on protein levels in main, age-adjusted models (x-axis) and in models additionally adjusted for *APOE4* carriage and years of education (y-axis).**

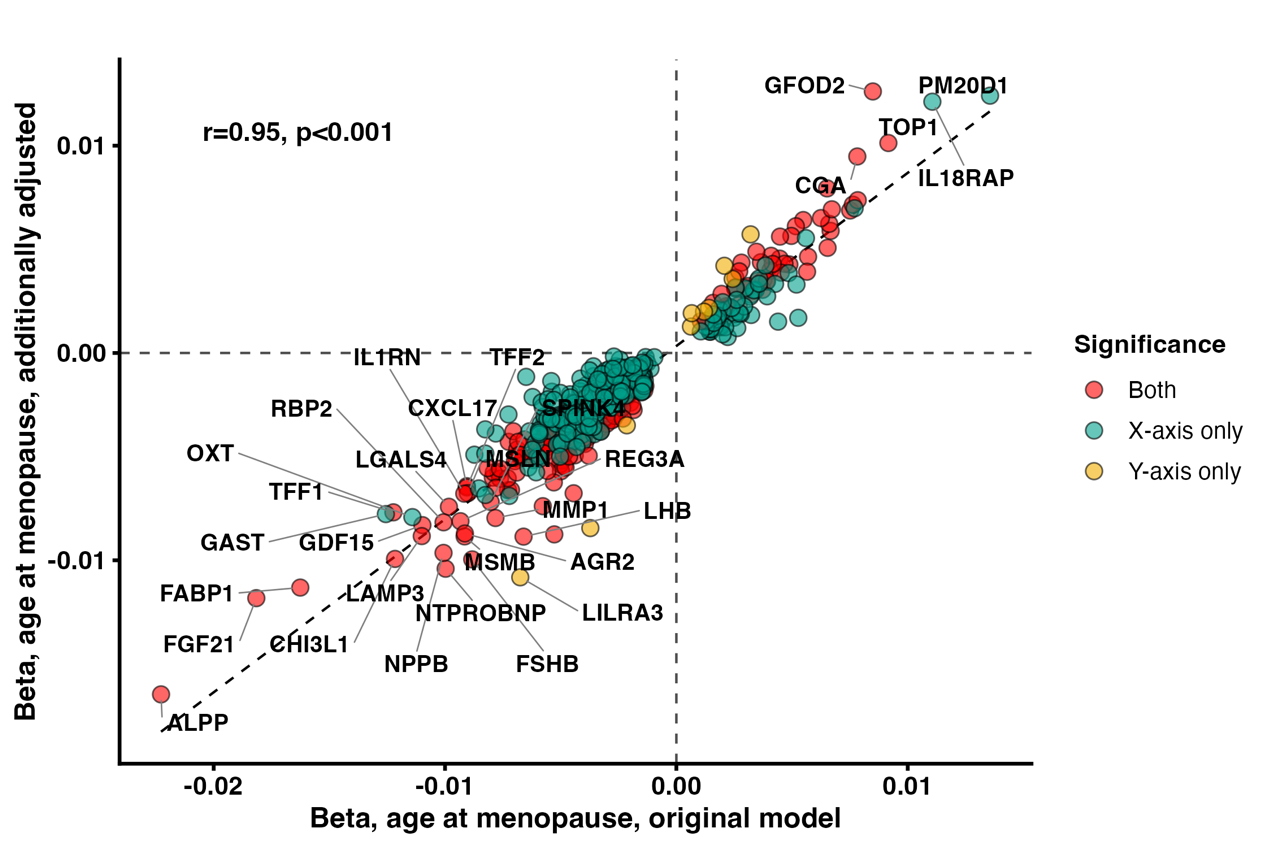

**Figure S8. Comparison of effect sizes of age at menopause on protein levels in women 50 years and older (x-axis) and in women 65 years and older (y-axis).**

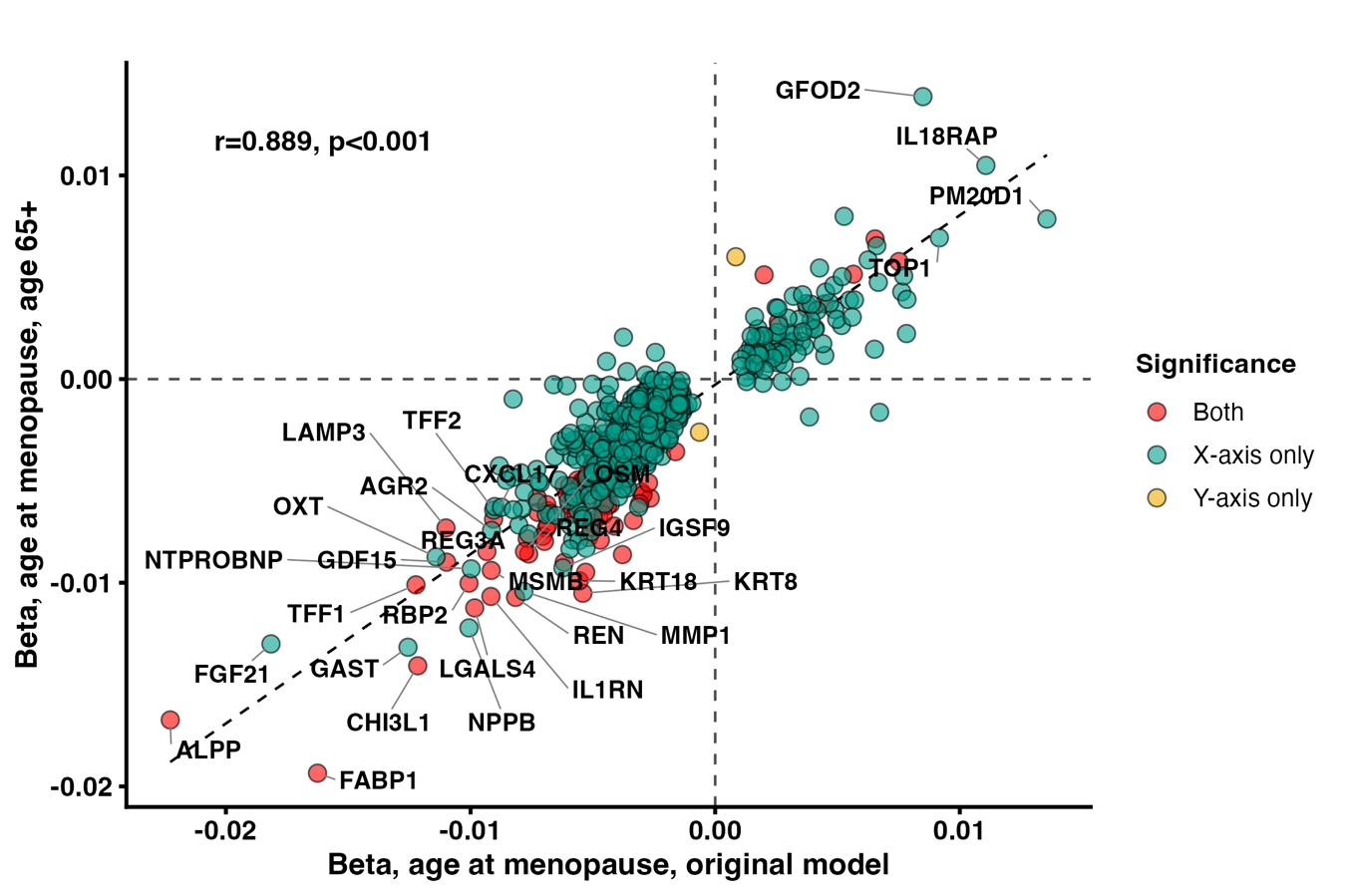

**Figure S9. Sex differences in associations of menopause timing proteins with dementia risk.**

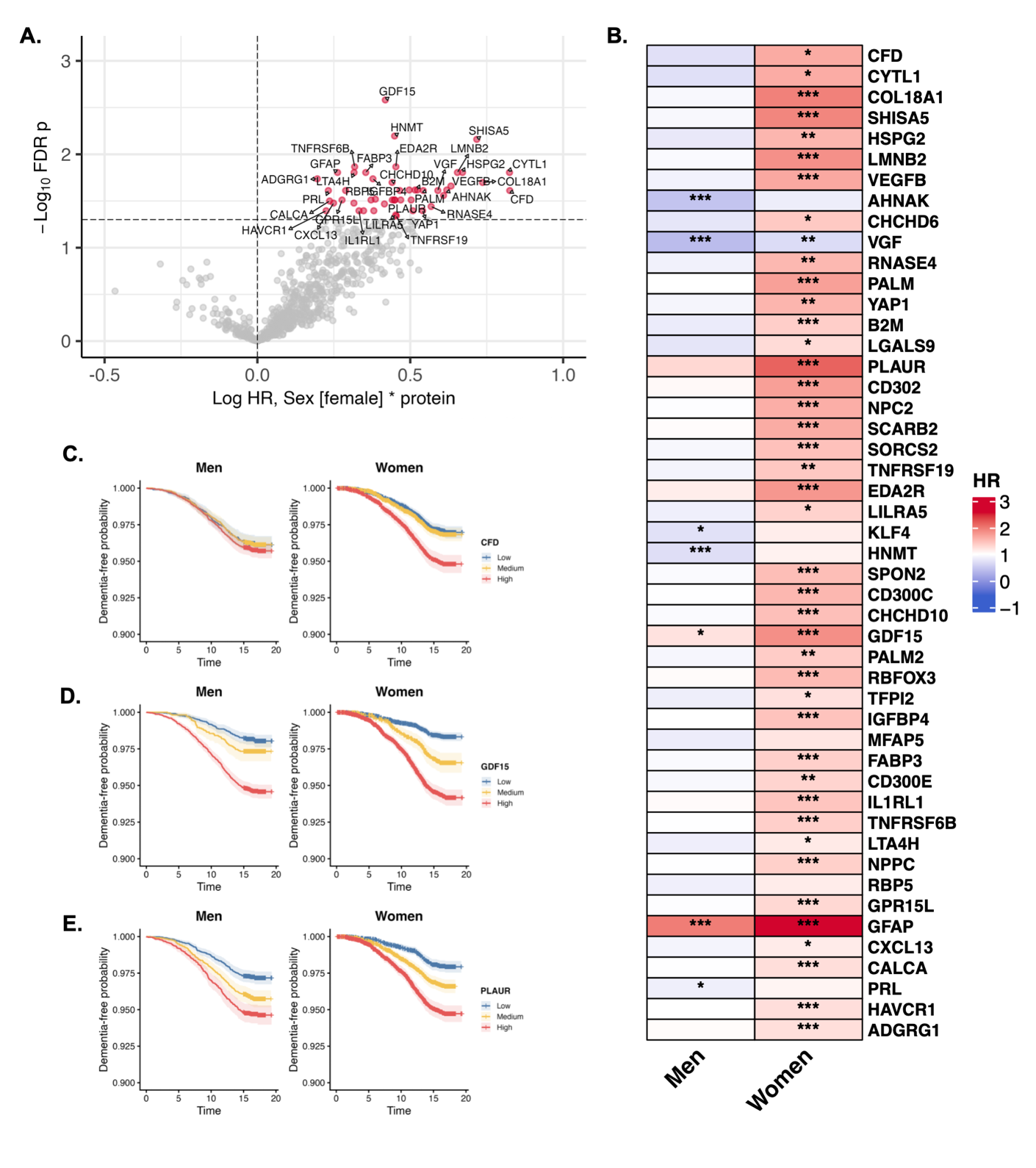

**(A)** Volcano plot showing effect sizes for interactions between menopause timing proteins and sex on incident all-cause dementia, from Cox proportional-hazards models adjusted for age. **(B)** Heatmap of sex-specific simple slopes from Cox proportional-hazards models testing interactions between menopause timing proteins and sex on incident all-cause dementia, adjusted for age. **(C-E)** Kaplan-Meier curves showing sex-stratified probabilities of progression to dementia, at lower, medium, and higher tertiles of **(C)** CFD, **(D)** GDF15, and **(E)** PLAUR.

**Figure S10. Menopause timing protein associations with total grey matter volume (GMV; scaled for head size), adjusted for age and time between Olink and MRI.**

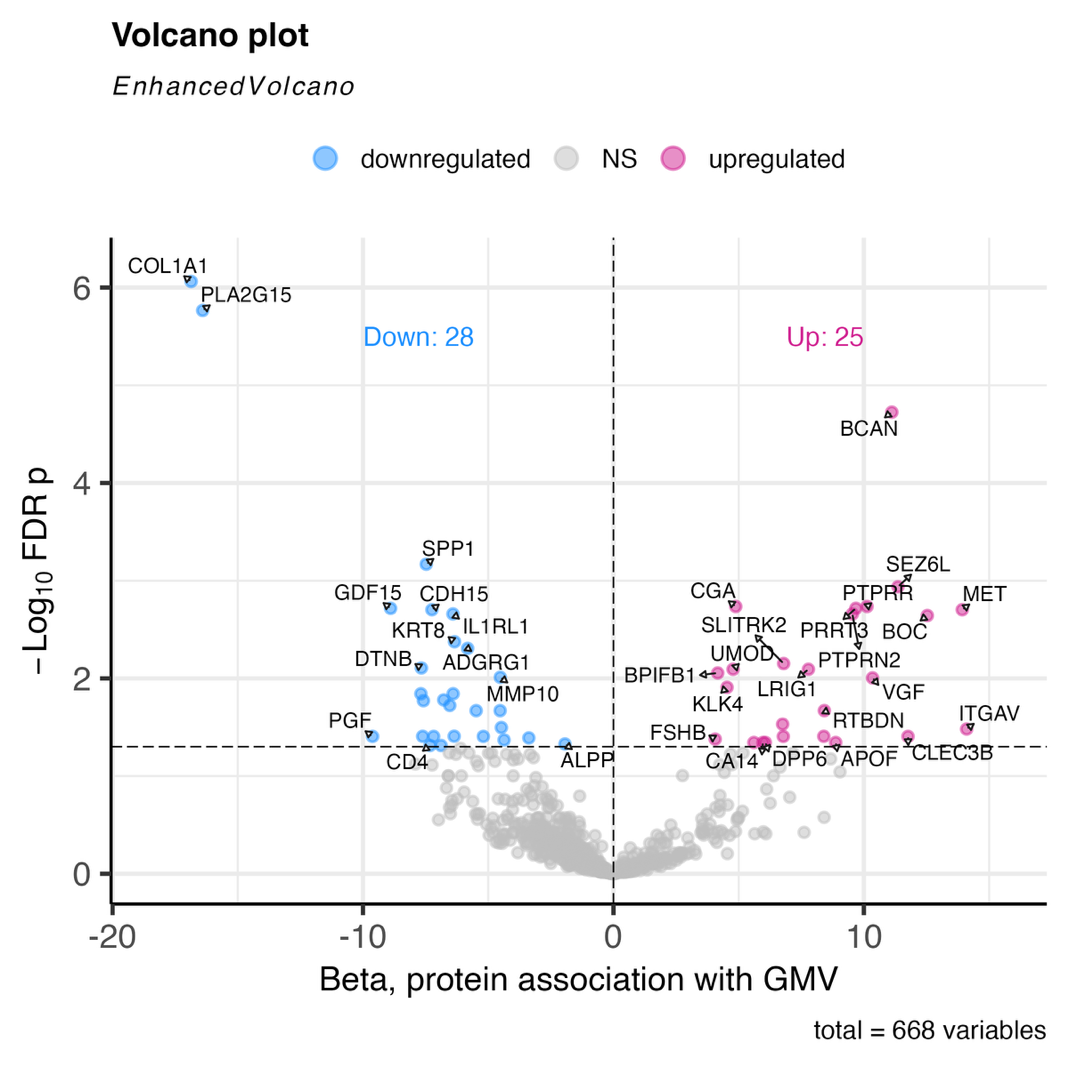

**Figure S11. Menopause timing protein associations with white matter hyperintensities (WMH; scaled for head size and log-transformed), adjusted for age and time between Olink and MRI.**

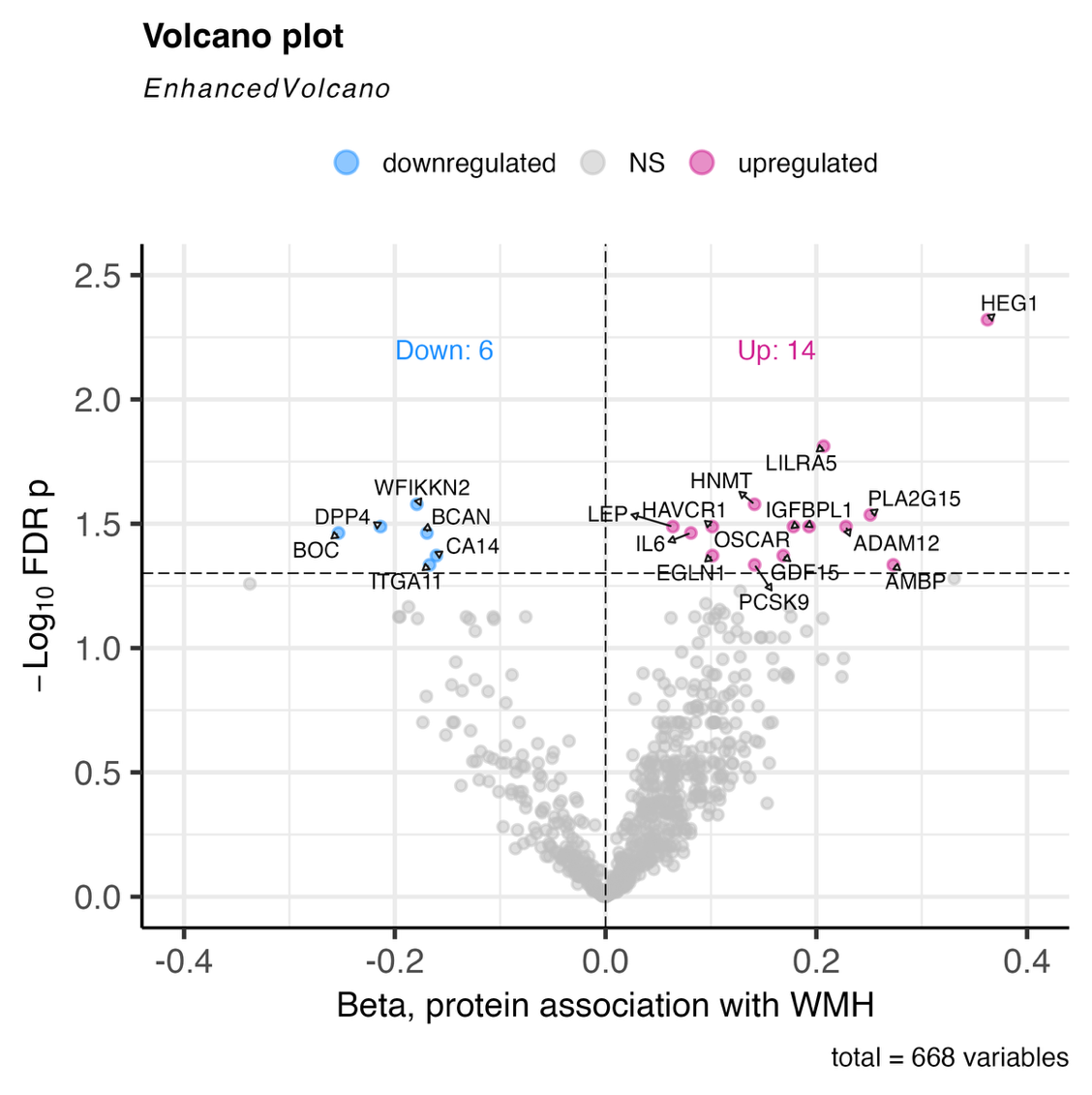

**Figure S12. Menopause timing protein associations with global fractional anisotropy (FA), adjusted for age and time between Olink and MRI.**

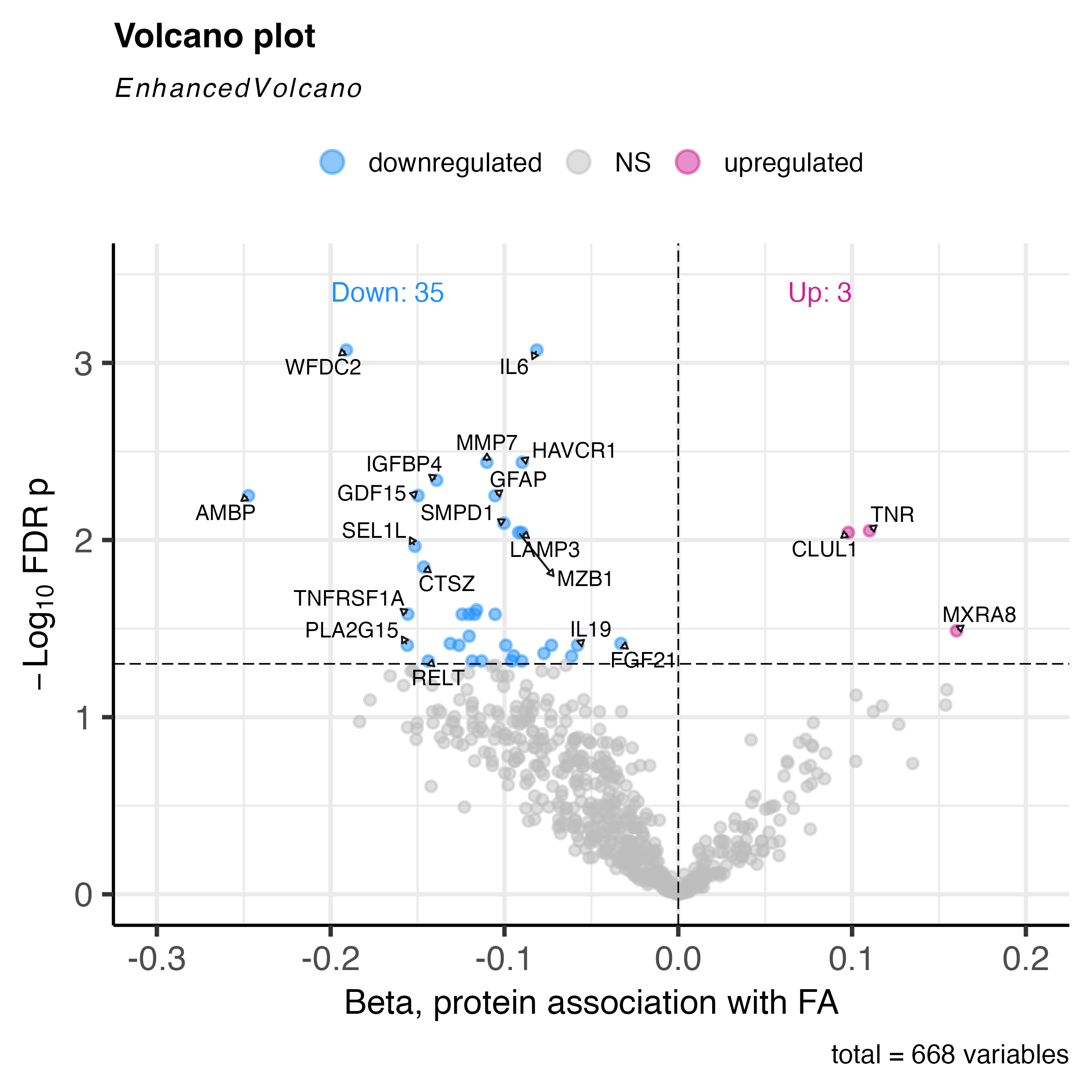

**Figure S13. Imputation evaluation metrics including distribution of ground truth vs. imputed mean average error (MAE) per protein in train and test splits (left) and total MAE rate in both train and test splits (right).**

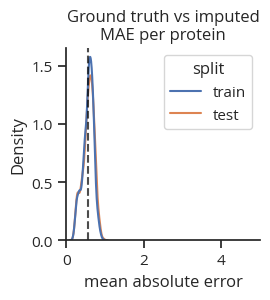

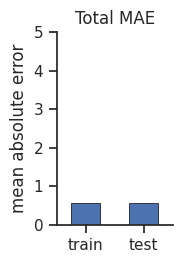
